# Surveillance of COVID-19 cases associated with dental settings using routine health data from the East of Scotland with a description of efforts to break chains of transmission from October 2020 to December 2021

**DOI:** 10.1101/2022.03.11.22272263

**Authors:** Niall Mc Goldrick, Emma O’Keefe

## Abstract

**Introduction:** Dental settings have been considered high risk setting s for COVID-19. A Dental Public Health Team in South East Scotland have worked to risk assess the situation timeously to break chains of transmission.

**Aim:** To present routine data produced from a contact tracing service for COVID-19 cases in the dental setting with a focus on transmission.

**Design:** Observational retrospective analysis of a routine data set of COVID-19 cases associated with a dental setting reported via the national contact tracing system for two health board areas in the east of Scotland.

**Methods:** COVID-19 cases were confirmed by PCR testing. Descriptive statistics are used to summarise the data collected over a 13-month period (Oct 2020-Dec 2021). A narrative presents themes identified during contact tracing that led to transmission within a dental setting and includes a case study.

**Results:** A total of 811 incidents are included. No evidence of staff to patient transmission or vice versa was found in this study. Staff to staff transmission occurred in non-clinical areas contributing to 33% of total staff cases.

**Conclusion:** Transmission of COVID-19 in a dental setting in the context of this study appears to be confined to non-clinical areas. Future pandemic plans should include tools to aid with implementation of guidance in non-clinical areas.

**In brief points:**

- Outbreaks of COVID-19 in a dental setting appear to be confined to the non-clinical areas of dental practices.
- We have found no evidence of staff to patient transmission or vice versa using our contact tracing methods.
- Future pandemic preparedness would benefit from including current quality improvement tools to aid with implementation of new standard operating procedures and other regularly changing guidance.

## Introduction

COVID-19 is a disease caused by the SARS-CoV-2 virus that was first detected in Wuhan, China in 2019 ^(1)^. On March 11^th^ 2020 the World Health Organisation (WHO) declared a global pandemic of COVID-19 ^(2)^. On March 23^rd^ the Chief Dental Officer for Scottish Government (CDO) ordered the closure of all general dental practices in Scotland and a retraction of dental services to Urgent Dental Care Centres (UDCC). On the same day a national lockdown with stay at home advice was issued by the Scottish Government. NHS Scotland established a national contact tracing programme ‘Test, Trace, Isolate and Support’ branded as ‘Test and Protect’ (TAP) for COVID-19 cases.

NHS Boards were asked to establish UDCCs throughout Scotland to provide urgent and emergency dental treatment. Urgent dental care was based on national guidance provided by Scottish Dental Clinical Effectiveness Programme published on 31/3/21 ^(3)^. General Dental Practitioners (GDPs) were asked to provide telephone triage for their registered patients and to provide a triage service for unregistered local patients. Restrictions on dental services eased in line with Scottish Government’s phased recovery and more treatment types became available^(4)^. In Phase 2, which began 22/6/21, and Phase 3, which began 13/7/21, of the route map to recovery, GDPs were remobilised to provide non aerosol generating procedures in local dental practices. As time went on routine dentistry reopened but with reduced foot fall due to ventilation issues and the need for continued physical distancing. NHS Scotland produced a National Dental Standard Operating Procedure that all NHS Scotland Dental Teams were required to follow when providing NHS dental care ^(5)^.

Throughout the pandemic TAP have identified cases of COVID-19, their contacts and the physical settings visited. Dental settings were identified as a discrete setting and required enhanced contact tracing due to use of various types of PPE and procedures performed.

The global literature on this topic is limited with study designs often restricted to cross sectional self reported outcome measures. One paper from Israel uses similar contact tracing methodology to that employed by this study ^(6)^. To the best of the authors knowledge there are no current UK studies presenting data on transmission of COVID-19 in a dental setting.

This paper aims to present observational routine data collected during the management of COVID-19 cases associated with dental settings in the East of Scotland with detail on transmission. Further, the paper presents the methods used to contact trace and manage COVID-19 cases associated with dental settings during the COVID-19 pandemic in Scotland.

This study was registered with NHS Fife’s Clinical Effectiveness Register and received approval as a service evaluation project.

### Objectives

- To present results from routinely collected data on COVID-19 cases associated with a dental setting including detail of onward transmission of SARS CoV-2 in a dental setting.
- To present methodology for risk assessing cases of COVID-19 associated with a dental setting.
- To highlight the areas in dental practices that give rise to onward transmission amongst staff with recommendations for improvement.

## Materials and methods

The study is observational in nature and is reported in line with the reporting of studies conducted using observational routinely collected data statement (RECROD) ^(7)^. We make use of routinely collected health data that has been accumulated by a dental public health (DPH) team during the management of COVID-19 cases associated with dental settings. The setting is two NHS Health Boards in the East of Scotland, NHS Fife and NHS Lothian. The population served by the Health Boards is circa 1.2 million people. There are 256 high street dental practices and two managed services operating across 30 sites.

A surveillance system was developed to capture all cases associated with dental settings in the two Health Board areas including patients, carers, and staff. There were no restrictions on participant characteristics and individual characteristics are not captured in the dataset. COVID-19 cases were identified by NHS Scotland’s national contact tracing programme ‘Test, Trace, Isolate and Support’ branded as ‘Test and Protect’ (TAP). Cases were confirmed following a positive PCR test for SARS CoV-2. Responsibility for the management of cases in high street dental practices was delegated by the Health Protection Teams in both Health Boards to the DPH Team. Responsibility for staff cases within managed dental services rested with the Occupational Health Team. Therefore, the data presented includes all cases reported in high street dental practices through the routes described. The dataset also includes all patient cases and some staff cases from managed dental services. The sample is a complete data set of all cases reported via the routes described. The observation period is 12/10/20 to 31/12/21. Data were collected in real time.

The data were available to the authors in the form of a routine data set with individuals marked by a coding system. The data were inspected and cleaned with emphasis on data completeness, removal of duplicates and uniformity of variables prior to analysis. Descriptive statistics were used to present the findings and all data were managed in MS Excel.

Following completion of initial contact tracing all cases associated with a dental setting in NHS Fife and NHS Lothian were referred to the dental public health team. Cases were defined as been associated with a dental setting if they were present there during their infectious period or if they were a member of staff. A detailed risk assessment of the case’s interaction within the dental setting was completed. Cases included patient, carers, and dental staff. All communication between DPH and TAP took place via email and cases were managed using NHS Scotland’s Case Management System (CMS), the national platform for recording all contact tracing activity for COVID-19 cases.

DPH carried out risk assessments for patients/carers and staff using the documents in supplementary material 1 and 2. The risk assessment process was agreed following meetings with colleagues in Health Protection, Infection Prevention Control and Occupational Health alongside guidance from Health Protection Scotland ^(8)^. The process was largely based on a risk assessment devised by pharmacy colleagues so that there was similarity across primary care ^(8)^. Risk assessment included detailed contact tracing, review of personal protective equipment used for patient facing care measured against the National Standard Operating Procedure, review of environmental cleaning following the presence of a known case and detailed review of the infection timeline for each case.

All cases were recorded on a central spreadsheet and updated as risk assessments progressed. The CMS was used to record the clinical notes related to the case and formal documentation of the risk assessment process. Contacts identified in dental settings were added to the CMS and notified using existing routes including SMS, email and in some cases by telephone.

Cluster investigations ensued where two cases were reported as having been associated with the same dental setting. Further detailed contact tracing and assessment of overlap of cases within the dental settings took place with a detailed look at staff and patient interaction. Transmission or an outbreak within dental settings was only confirmed following this detailed contact tracing and where two or more cases could be linked ^(8)^. Contacts of a confirmed case who were exposed in a dental setting were captured in ongoing surveillance and if contacts went onto develop COVID-19 or tested positive on PCR highlighted a chain of transmission was established.

Action following risk assessment often included escalation to Health Protection of complex cases or outbreaks requiring specialist health protection input. An Incident Management Team or Problem Assessment group could be established to work through risk assessments and more in-depth investigation of clusters/outbreaks. Most cases were managed by DPH with verbal or written advice provided to dental teams and formal isolation advice issued to cases or contacts. Advice included reference to environmental cleaning as per the National Infection Prevention Control Manual and NHS Scotland National Standard Operating Procedure for Dental Teams ^(5)^. DPH colleagues could recommend dental practices consider the NHS Education for Scotland Quality Improvement in Practice Training Team in Dental Infection Prevention Control for bespoke and in-depth sessions as required. Constant reflective practice was employed throughout with frequent team meetings to enable sharing, learning and improvement for both the dental settings and the DPH response.

## Results

A total of 811 incidents associated with dental settings were managed in the time period 12/10/20-31/12/21. Fifty-nine incidents required advice but did not require a full risk assessment due to a variety of factors e.g., staff cases not been present in the workplace during their infectious period. Of the remainder, 250 cases were identified as dental staff. Five hundred and two cases were identified as patients and carers. Figure 1 below presents staff and patient cases mapped against the cases in the general Scottish population overtime.

**Figure 1:**
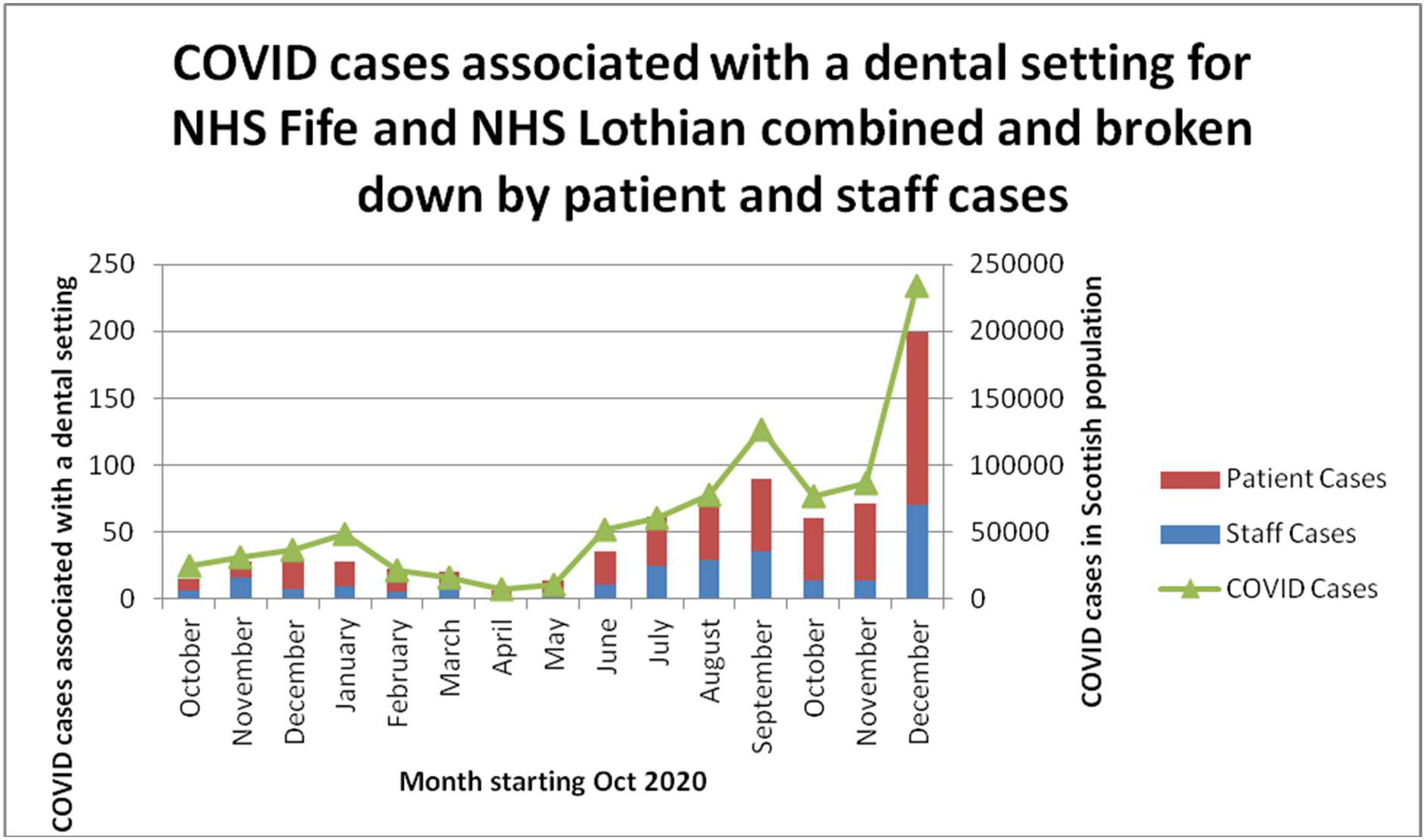
COVID-19 cases (patient and staff) associated with a dental setting for NHS Fife and NHS Lothian and cases in the general Scottish population for time period October 2020-December 2021 mapped with cases in the general population.

The breakdown of cases for the entirety of the observation period was 31% staff and 69% patients.

In this cohort of cases there has been no evidence of transmission from patients to staff or staff to patients. There were 20 instants of staff-to-staff transmission resulting in outbreaks during the observation period. This totalled 83 cases representing 33% of total recorded staff cases and generating 31 close contacts amongst remaining staff. The range of staff cases in an outbreak varied from two to seven cases.

There were three instants of transmission amongst staff at social events outside the dental setting amongst staff amounting to 10 cases.

Embedded below is a case study taken from within this dataset to demonstrate how COVID-19 can spread rapidly through a practice and some of the common human factors that can contribute to transmission. In this case study six staff members were confirmed COVID-19 cases and an additional four staff were identified as contacts. The outbreak significantly impacted the ability of the dental practice to deliver patient care resulting in emergency measures to provide care for patients. This type of situation could affect the financial security of a practice and potentially impact on reputational risk. Figure 2 below presents results from the contact tracing tool used to map this outbreak.

**Figure 2:**
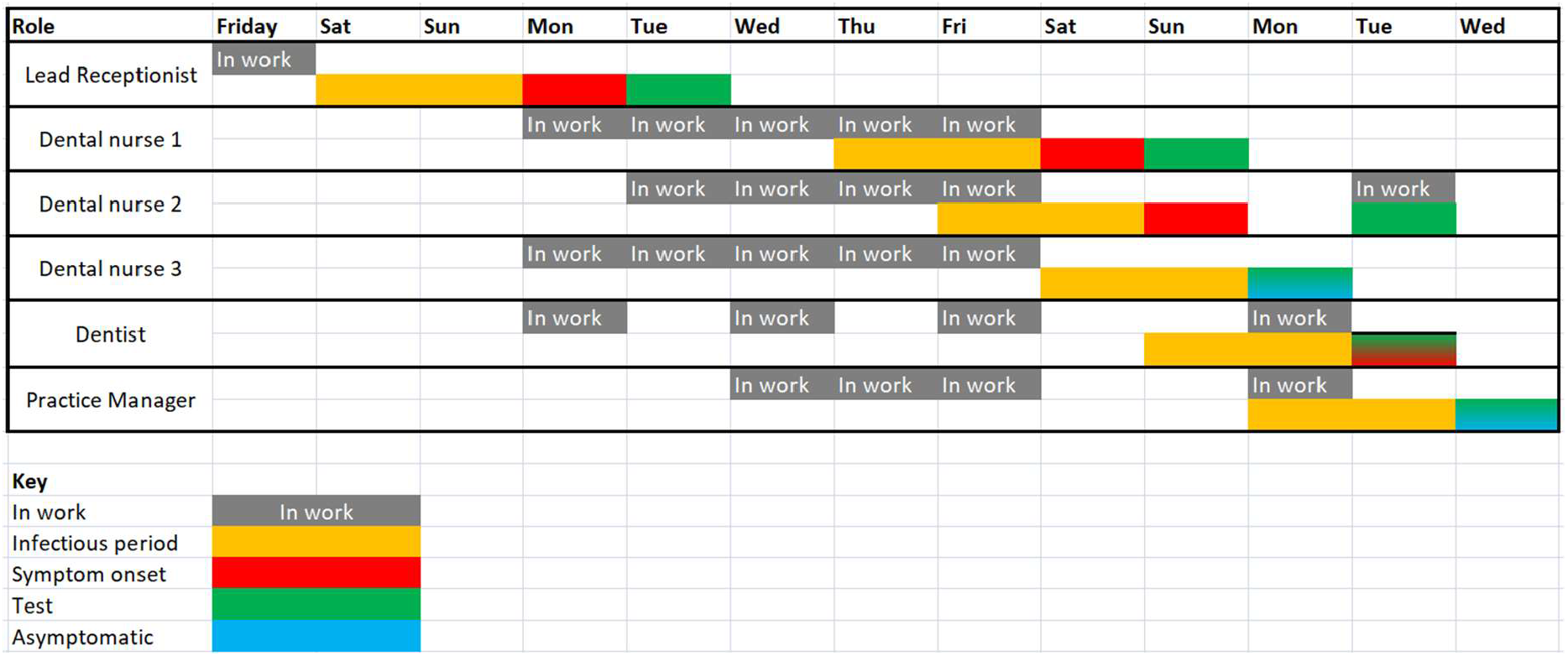
Contact tracing timeline for cases associated with one dental practice aiding with assessment for possible routes of transmission.

Prior to and unrelated to this outbreak, the practice had an isolated case of COVID-19 amongst staff. At that time the practice declared having a risk assessment and standard operating procedure in place that had been reviewed and no further action was required. Despite these processes being recorded the outbreak occurred two weeks later.

Several key areas for improvement were identified:

### Car sharing

Two members of staff shared a car journey without precautionary measures were taken such as wearing face coverings, distancing as best possible and keeping windows open.

### Staff changing room

Members of staff utilised a small unventilated changing room at the same time with more people than recommended for the space.

### Kitchen area

this area was an internal room, with no windows or ventilation. Environmental cleaning did not take into account shared use items and surface cleaning.

### Sharing office space

due to IT issues multiple staff shared an office space, recommended for fewer people, for prolonged periods in a poorly ventilated room.

### Staff health and wellbeing

one staff member presented at work while feeling unwell to fulfil their work duties and support service delivery.

Table 1 presents common themes from outbreaks that may have contributed to transmission.

**Table 1:**
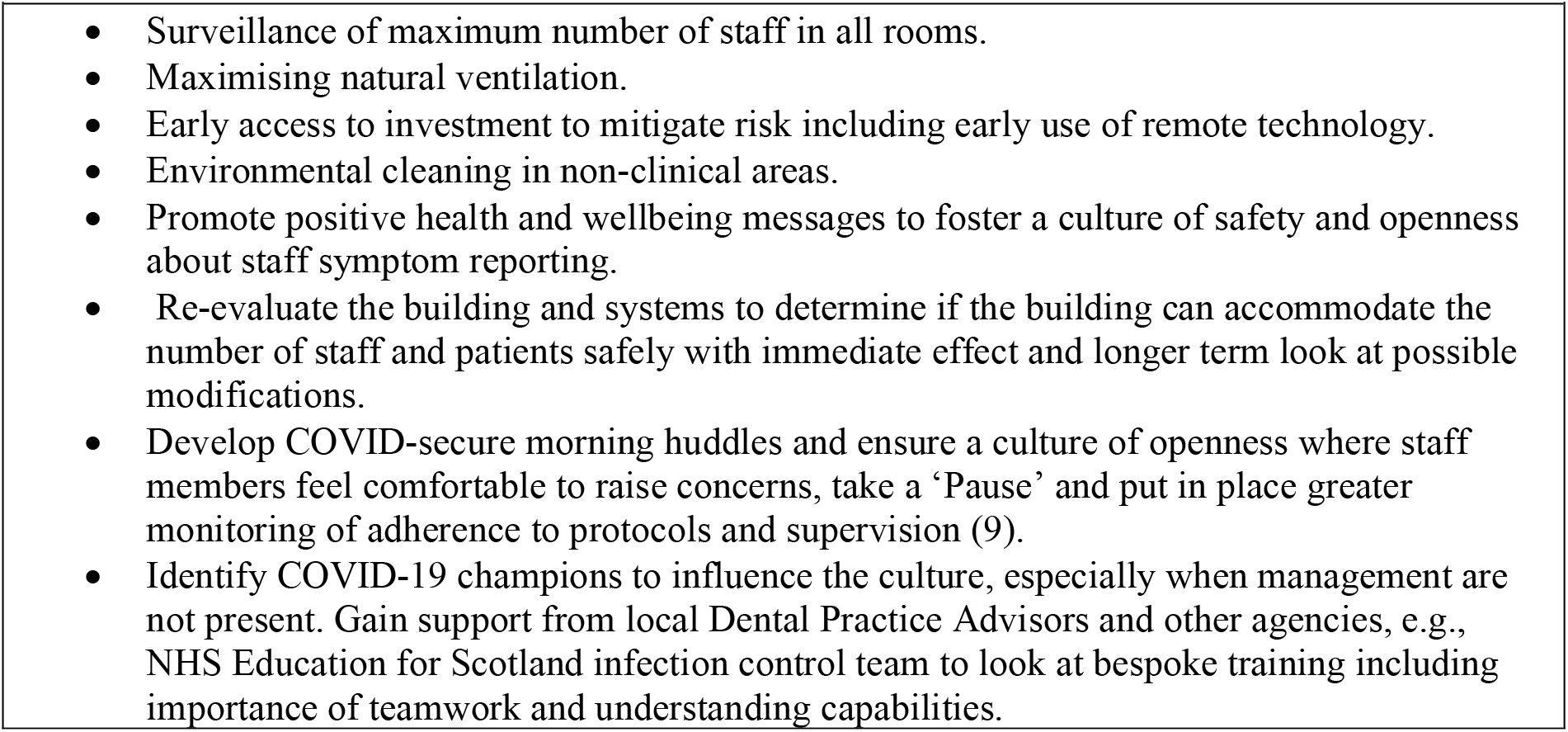
Common themes identified as contributing to transmission during outbreaks to be considered in future pandemic preparedness.

## Discussion

The trend of cases associated with dental settings follows the trend in the general population. Throughout the observation period no evidence of transmission between staff and patients or vice versa was found. This is following in depth contact tracing and risk assessment with some early cases having thorough investigation by multi-disciplinary Incident Management Teams with specialist health protection and infection prevention control input. Previous work in Scotland has demonstrated a low prevalence of COVID-19 (0.5%) amongst dental patients who are screened and are asymptomatic ^(10)^. Considering the findings of this study in tandem with Conway et al’s work presents the suggestion of a low risk of occupational exposure for dentists from patients in the context of the infection prevention control measures at the time.

However, transmission between staff was demonstrated multiple times in non clinical areas. Despite having appropriate risk assessments and protocols in place this did not always translate to staff behaviour and action. Quality improvement tools and learning from the Scottish Patient Safety Programme were lacking in the implementation of protocols. There was a reliance on staff reading, signing for, and hopefully implementing ever changing protocols. In future pandemics with a similar virus guidance and implementation methodology should take into account findings presented in table 1.

Strengths of this study include how the DPH team received notification of all cases of COVID-19 associated with dental settings in the area. It was possible to keep continued surveillance for onward transmission in dental settings therefore providing a good level of ascertainment of cases associated with dental settings. Collaboration between HPT, DPH and primary care provided a holistic response to cases in dental settings pulling on local experience and knowledge of dental provision in the area.

Limitations include the fact that genome sequencing was not employed in contact tracing for any of these cases. Cases that did not engage with the national contact tracing system and those that did not report an interaction with a dental setting will not have been picked up. Contact tracing also relies on recall of individuals and for cases to be well enough to take part. Some cases experiencing the worst symptoms and morbidity associated with COVID-19 will not have been able to partake fully or at all in contact tracing reducing ability to break chains of transmission. Cluster and outbreak locations were not visited by DPH to assess the measures in place but instead relied upon what was reported by local management teams.

The DPH team took the opportunity to constantly reflect using regular team huddles to debrief, share learning and ask questions. Reactive trouble shooting huddles took place via MS TEAMS daily. Use of a shared folder for documenting actions on cases and maintaining case notes provided ease of handover between staff working remotely. The DPH team were well place due to existing networks that straddle both public health and dentistry.

The findings from this paper would likely be comparable with other areas of Scotland or the UK where similar contact tracing processes were in place.

## Conclusion(s)

We have found no evidence of staff to patient or patient to staff transmission in this study. Staff to staff transmission within dental setting was a repeated problem suggesting that more efforts to implement and continually reiterate preventive guidance in non-clinical areas are required in dental settings during a pandemic.

## Data Availability

The raw data set is not available publicly.

## Declaration of interests

The authors declare no conflicts of interest.

## Author contributions statements

Both authors have contributed equally to this study and should be viewed as joint first authors.

## Acknowledgements

We wish to acknowledge Jacqueline Burns, Specialty Registrar in Dental Public Health in NHS Fife/Lothian was a key member of the team handling situations and took as similar approach to NHS A&A in her role as Consultant in Dental Public Health. We are grateful to our Health Protection Team and Test and Protect colleagues for supporting us. We are grateful to the practice involved in the embedded case study for allowing us to share their challenging experience.

## Notes

### Competing Interest Statement

The authors have declared no competing interest.

### Funding Statement

This study did not receive any funding.

### Author Declarations

The Research and Development Department of NHS Fife waived ethical approval for this work on the basis that it is service evaluation. The project has been registered and reviewed internally as part of clinical effectiveness activity.

