## Supplementary material 1 for "Surveillance of COVID-19 cases associated with dental settings using routine health data from the East of Scotland with a description of efforts to break chains of transmission from October 2020 to December 2021"

**Background information required for Dental Public Health Team for confirmed positive patient in a dental setting**

**This information will assist the Dental Public Health team (DPH) with their risk assessment of the dental setting.**

It is the responsibility of Test and Protect to gather information directly from the person who tested positive to identify household and close contacts in the community. The role of the Dental Public Health team is to liaise with the dental setting to risk assess and identify any further contacts in that environment and to review control measures required to prevent further spread. We have found the information provided is invaluable in this process.

**Definition of Infectious period:** the infectious period is defined from 48hrs prior to the onset of symptoms until 10 days after symptom onset.

**Infectious period of the case:**

**Definition of a 'close contact':** close contact is defined as having a person having evidence of the following, during the infective period:

1. ***Direct contact:***

• face to face contact with a case within 1 metre for any length of time, including:

- being coughed on
- having a face-to-face conversation
- having skin-to-skin physical contact

• any contact within 1 metre for one minute or longer without face-to-face contact

• a person who has travelled in a small vehicle with someone who has tested positive for coronavirus (COVID-19) or in a large vehicle near someone who has tested positive for coronavirus (COVID-19).

2. ***Proximity contact:***

• A person who has been between 1 and 2 metres of someone who has tested positive for coronavirus (COVID-19), for more than 15 minutes cumulatively during period 48 hours prior to and 10 days after the case's symptom onset

**Please note** the above criteria are irrespective of wearing a face covering as face coverings are non-medical grade coverings they do not exempt the wearer from being considered a close contact if physical distancing is not maintained with an individual who tests positive.

**Fluid resistant surgical masks (Type IIR)** are medical grade masks and protect the wearer. As a result, they *do* exempt the wearer from being considered a close contact if physical distancing is not maintained. However, this is in the context of health care settings or settings associated with healthcare and only with proper training on the use of fluid resistant surgical masks and other personal protective equipment (i.e. gloves, aprons, eye protection). Ineffective or improper use of fluid resistant surgical masks and/or breaches in personal protective equipment will not adequately protect the wearer and so the wearer will then be considered a contact if physical distancing is not maintained with an individual who tests positive, and they meet the definition of a contact.

| | | | | |
|---|---|---|---|---|
| **Risk Assessment checklist for positive patient in dental setting Clinic Location:** | | | | |
| **Date attended dental setting:** | | | | |
| **Names of dental team members providing care:** | | | | |
| **Patient Name:** | | | | |
| **Case ID (CMS number):** | | | | |
| **Infectious period of case:** | | | | |
| Contact Traced: | Consent to discuss case: | | | |
| Type of appointment: | **AGP** | | **Non-AGP** | |
| Length of appointment: | | | | |
| Does the practice have a SOP? | **YES** | | **No** | |
| Does it follow the National SOP? (if not please specify what guiding principles it follows) | | | | |
| When was it last updated? | | | | |
| How are staff aware and adhering to SOP?<br><br>e.g. read/signed/huddle/practice meeting | | | | |
| COVID screening questionnaire:<br><br>COVID screening checked at time of appointment?<br><br>Entered in patient record: | **Y/N**<br><br>**Y/N**<br><br>**Y/N** | | | |
| PPE appropriate: | **FRSM/IIR** | **Visor** | **Gloves** | **Apron** |
| | **FFP3** | **Visor** | **Gloves** | **Gown** |
| If FFP3 worn | **NHS** | **Reusable** | **SOPs** | |
| Patient Flow (describe how this works in the | | | | |

| | | | |
|---|---|---|---|
| dental setting) : | | | |
| Is waiting room in operation: | | | |
| Payment mechanism: | **Cash** | **Contactless** | **Card** |
| Cleaning (Environmental cleaning including type of wipes used (if applicable) and patient journey): | | | |
| What PPE is used for cleaning: | **FRSM** | **Gloves** | **Apron** |
| Contacts within dental setting identified: Y/N<br><br>(Please use the information on the previous pages to identify possible contacts) | If yes provide detail: | | |
| Are staff involved in regular (twice weekly) asymptomatic Lateral Flow Device Tests (LFDs)? | Yes/No/Some | | |
| Have all staff received their COVID vaccinations? | Y/N | | |
| Other: | | | |
| Actions: | | | |
| Risk assessment completed by:<br><br>Date: | | | |

FOR DENTAL PUBLIC HEALTH USE: SOPs, Risk Assessment checked: Y/N

SUMMARY FOR CMS:
