## Supplementary material 2 for "Surveillance of COVID-19 cases associated with dental settings using routine health data from the East of Scotland with a description of efforts to break chains of transmission from October 2020 to December 2021"

### Background information required for Dental Public Health Team for confirmed positive case in dental workplace setting

**This information will assist the Dental Public Health team (DPH) with their risk assessment and identification of close contacts of the case in the dental workplace setting who will be advised to self isolate as a precautionary measure to stop the spread of COVID-19**

It is the responsibility of Test and Protect to gather information directly from the person who tested positive to identify household and close contacts in the community. The role of the Dental Public Health team is to liaise with the dental workplace to identify further contacts in the workplace environment and consider control measures required to prevent further spread. We have found the information workplaces provide is invaluable in this process.

Office use: Risk assessment
SOP
Declaration

| | | |
|---|---|---|
| Workplace Name | | |
| Name of manager/key contact and contact details (phone number/email) | | |
| Name of staff member tested positive and contact details | | |
| Type of test (Please tick next to the appropriate test(s) | **LFT:** | **PCR:** |
| Role of staff member | | |
| Whole time equivalent unit | | |
| When did the positive staff member's symptoms start? | | |
| When were they last in the workplace? | | |
| Are there any known or suspected Covid cases in patients or colleagues? | | |
| What dates did the case work during their infectious period? See Page 1 for the infectious period | | |
| Please provide a brief overview of a typical day in the workplace for the staff member who has tested positive | | |
| Please provide a brief overview of control measures that are in place in the workplace to keep both staff and members of the public safe. Please also state if your standard operating procedure conforms with the NHS Scotland National SOP. | **Include info Don/Doff, IPC, environmental cleaning please reference any guidance referred to.** | |
| Please provide a brief overview of any risk assessment you have undertaken in the workplace already? *(e.g. temporary closure, communication to staff team, deep cleaning etc)* | | |
| Specify what role the case had on the days worked (during infectious period to last day worked) | **E.g. chairside, LDU, on reception** | |
| Using the definitions provided on page 1, do you think any members of the public would fulfil the definition of a close contact of the staff member who has tested positive?<br><br>*If yes, please discuss this with the Dental Public Health Team and collate any lists of these members of public that you have (names and contact details)* | **Names and contact details of members of public who may be contacts of the case from contact at the workplace:**<br>**+ type of treatment patient had and length of the appointment + DON/DOFF** | |
| Did the staff member use any communal areas – staff room/changing room etc including handover/breaks/meetings? | **Key areas to consider for staff members who may have had contact with the case:**<br>• **Coffee breaks/break rooms**<br>• **Colleagues who sit or work near the case** | |

Office use: Risk assessment
SOP
Declaration

| | |
|---|---|
| Using the definitions provided on page 1, do you think any members of your staff team would fulfil the definition of a close contact of the staff member who has tested positive?<br><br>*If yes, please inform these staff that they are close contacts of the case (without disclosing the name of the staff member) and that they should self-isolate for 14-days from date of last contact with the case. The Dental Public Health Team are happy to provide guidance on this if you are unsure about the correct dates to use. Please provide their details in table 2.* | • **Smoking breaks**<br>• **Colleagues who travel together**<br>• **Kitchen/lunch rooms**<br>• **Meetings/meeting rooms**<br>• **Car sharing** |
| Do you have any staff members who work on different sites and who may have had close contact with the case (as per the definition on page 1) | **Please record their names and contact details in Table 2 and alert the DPH Team** |
| Could there have been any other visitors to the workplace (e.g. other professionals, work/repairs teams, delivery people) who may have had close contact with the case (as per definition on page 1) | **Please record their names and contact details in Table 2 and alert the DPH Team** |
| Do you have any other staff members who are confirmed positive cases or who are symptomatic?<br><br>*If yes, please provide their names* | **Names and contact details of other staff members who have tested positive or who are symptomatic:** |
| Are staff taking part in lateral flow device testing? If yes, how often? | |
| Is there any link between staff members who have tested positive or who are currently symptomatic? | |
| Have all staff received COVID vaccinations? | **Yes/No** |

**Table 2:**

**Please record all staff members who you have identified as close contacts of the case in the workplace setting (if they have not already been contacted by Test and Protect):**

| **Full Name** | **Contact number** | **Nature of contact (e.g. works beside case or contact in coffee room)** | **Date last in contact with confirmed positive case** | **Advised self isolation dates*** | **Job role** | **WTE**** |
|---|---|---|---|---|---|---|

Office use: Risk assessment
SOP
Declaration

*** Self-isolation period is 10-days from day of last contact with the person who has tested positive – please ask the Dental Public Health team if you need any clarification on what date to advise.**

****WTE: whole time equivalent working i.e 0.5 for 50% week, 1 for 100%**

**Please note, a negative test does not exempt a close contact from having to complete their full 10 day isolation period.**

Office use: Risk assessment
SOP
Declaration
